# Pandemic dynamics of COVID-19 using epidemic stage, instantaneous reproductive number and pathogen genome identity (GENI) score: modeling molecular epidemiology

**DOI:** 10.1101/2020.03.17.20037481

**Authors:** DJ Darwin R. Bandoy, Bart C. Weimer

## Abstract

**Background:** Global spread of COVID-19 created an unprecedented infectious disease crisis that progressed to a pandemic with >180,000 cases in >100 countries. Reproductive number (R) is an outbreak metric estimating the transmission of a pathogen. Initial R values were published based on the early outbreak in China with limited number of cases with whole genome sequencing. Initial comparisons failed to show a direct relationship viral genomic diversity and epidemic severity was not established for SARS-Cov-2.

**Methods:** Each country’s COVID-19 outbreak status was classified according to epicurve stage (index, takeoff, exponential, decline). Instantaneous R estimates (Wallinga and Teunis method) with a short and standard serial interval examined asymptomatic spread. Whole genome sequences were used to quantify the pathogen genome identity score that were used to estimate transmission time and epicurve stage. Transmission time was estimated based on evolutionary rate of 2 mutations/month.

**Findings:** The country-specific R revealed variable infection dynamics between and within outbreak stages. Outside China, R estimates revealed propagating epidemics poised to move into the takeoff and exponential stages. Population density and local temperatures had variable relationship to the outbreaks. GENI scores differentiated countries in index stage with cryptic transmission. Integration of incidence data with genome variation directly increases in cases with increased genome variation.

**Interpretation:** R was dynamic for each country and during the outbreak stage. Integrating the outbreak dynamic, dynamic R, and genome variation found a direct association between cases and genome variation. Synergistically, GENI provides an evidence-based transmission metric that can be determined by sequencing the virus from each case. We calculated an instantaneous country-specific R at different stages of outbreaks and formulated a novel metric for infection dynamics using viral genome sequences to capture gaps in untraceable transmission. Integrating epidemiology with genome sequencing allows evidence-based dynamic disease outbreak tracking with predictive evidence.

**Funding:** Philippine California Advanced Research Institute (Quezon City, Philippines) and the Weimer laboratory.

**Research in context:** Reproductive number is (R) an epidemiological parameter that defines outbreak transmission dynamics. While early estimates of R exist for COVID-19, the sample size is relatively small (<2000 individuals) taken during the early stages of the disease in China. The outbreak is now a pandemic and a more comprehensive assessment is needed to guide public health efforts in making informed decisions to control regional outbreaks. Commonly, R is computed using a sliding window approach, hence assessment of impact of intervention is more difficult to estimate and often underestimates the dynamic nature of R as the outbreak progresses and expands to different regions of the world. Parallel to epidemiological metrics, pathogen whole genome sequencing is being used to infer transmission dynamics. Viral genome analysis requires expert knowledge in understanding viral genomics that can be integrated with the rapid responses needed for public health to advance outbreak mitigation. This study establishes integrative approaches of genome sequencing with established epidemiological outbreak metrics to provide an easily understandable estimate of transmission dynamics aimed at public health response using evidence-based estimates.

**Added value of this study:** Estimates of R are dynamic within the progression of the epidemic curve. Using the framework defined in this study with dynamic estimates of R specific to each epicurve stage combined with whole genome sequencing led to creation of a novel metric called GENI (pathogen genome identity) that provides genomic evolution and variation in the context of the outbreak dynamics. The GENI scores were directly linked and proportional to outbreak changes when using disease incidence from epicurve stages (index, takeoff, exponential, and decline). By simulating short and standard (2 day and 7 day, respectively) serial intervals, we calculated instantaneous R followed by a global comparison that was associated with changes in GENI. This approach quantified R values that are impacted by public health intervention to change the outbreak trajectory and were linked to case incidence (i.e. exponential expansion or decelerating) by country. Integrating viral whole genome sequences to estimate GENI we were able to infer circulation time, local transmission, and index case introduction. Systematic integration of viral whole genome sequences with epidemiological parameters resulted in a simplified approach in assessing the status of outbreak that facilitates decisions using evidence from genomics and epidemiology in combination.

**Implications of all the available evidence:** This study created a framework of evidence-based intervention by integrating whole genome sequencing and epidemiology during the COVID-19 pandemic. Calculating instantaneous R at different stages of the epicurve for different countries provided an evidence-based assessment of control measures as well as the underlying genomic variation globally that changed the outbreak trajectory for all countries examined. Use of the GENI score translates sequencing data into a public health metric that can be directly integrated in epidemiology for outbreak intervention and global preparedness systems.

## Introduction

Outbreaks are defined by the reproductive number (R)^1,2^ a common measure of transmission. Probability of further disease spread is evaluated based on the threshold value with likely expansion for values >2 and decline with values of <1. R is the main component for computing the needed proportion of the population to be vaccinated based on herd immunity^3^. The expansion of COVID-19 was determined with the earliest estimate of R = 2.2 (95% CI, 1.4 to 3.9) using serial intervals for 424 patients in Wuhan, China^4^. Recalculation with 2033 cases estimated R = 2.2 to 3.6^5^. However, estimates of R for other countries where cases were found as the outbreak grew in China were not done routinely and currently a fixed estimate R is used based on the refined estimate from China. However, this is falling short in predicting the spread of the pandemic and expansion within individual locations, suggesting that R is not likely to be constant and likely to be dynamic for each outbreak location that results in underestimates of the spread rate. This limitation is hindering epidemic dynamics as previously noted due to the parameter is context specific and dynamic^1,2^. Hence, there is a need to rapidly estimate country specific R values during the epidemic. This will provide global comparisons of expansion at each location.

The Wallinga and Teunis method for R estimation requires input of outbreak incidences and the serial interval (i.e. the period between the manifestation of symptoms in the primary case and the onset of symptoms in secondary cases)^6^. This approach was implemented in a web resource to estimate R during epidemics^7^. A key advantage of the approach is the ease of production of credible intervals compared to other maximum likelihood estimation approaches. Yet to be done is integration of viral genetic variation with R estimates but one study found that there was no obvious relationship between R, severity of the epidemic and COVID-19 genome diversity^20^.

COVID-19 has reached global spread in all continents except Antarctica and was defined to be a pandemic by the World Health Organization (WHO) in March 2020^8-10^. The outbreak dynamics are different between countries as well as varying within individual countries. In part this is due to varying and diverse healthcare systems, socio-cultural contexts, and rigorous testing. Considering the lack of containment globally, except in Singapore, Hong Kong, and Taiwan, we hypothesized that previously calculated R values do not provide reliable estimates because they are more dynamic than is being considered and that influx of new cases and viral mutation are likely sustaining expansion. While viral sequencing is occurring, it is not being effectively integrated with epidemiological information because there is no existing framework for that to systematically occur.

In spite of no clear path for deep integration of viral variation the current pandemic has demonstrated the public health unity for sharing COVID-19 whole genome sequences with an unprecedented openness. By quickly sharing the genome sequences it enables investigation of the genome variation during the outbreak using multiple approaches and samples of the virus genome space. It is approaching a viral population scale, which provides additional information that cannot be gleaned with few sequences. Prior work established the value of estimating transmission dynamics of rapidly evolving RNA viruses and highlights the capability to infer transmission during outbreaks coupled with pathogen genomes^11,12^. This approach was validated in EBOV and MERS. Each virus variant is separated by only several mutations yet produces new dynamics during the outbreak^13,14^. Rapidly evolving pathogens undergo genome sequence mutation, selection pressure, random drift and stochastic events between infected individuals^11^. Even small changes in the genome enable transmission that is determined by accounting for the mutations between isolate sequences. It is recognized that the COVID-19 genome is changing over the outbreak but there is controversy about the impact and specifics of the exact mutations. In this study, we used incidence data to derive R and compared country specific COVID-19 infection dynamics with viral population genome diversity. By incorporating R, epidemic curve timing, and viral genome diversity we created a systematic framework that deduced how viral genome diversity can be used to describe epidemiological features of an outbreak before new cases were observed. This was done by creating a genome diversity metric that was directly and systematic integrated to provide context and allowed quantification of the infection dynamics globally that are divergent from the early estimates with genomic evidence. We call this approach pathogen genome identity (GENI) scoring system. Using GENI differentiated each stage of the outbreak. It also indicated cryptic local transmission from surveillance systems. This a defining advantage of using sequences as previous cryptic transmission can be inferred in the genomic sequences.

## Methods

Incidence data is based on daily Chinese CDC and WHO situations reports as compiled by the Center for Systems Science and Engineering (CSSE) by the John Hopkins University (Baltimore, MD, USA) that was accessed on March 1, 2020^15^. We constructed epidemic curves or epicurves from the incidence data and classified country status accordingly. We defined four groups that characterize increasing expansion with a decline phase.

The extracted time series case data served as the input for determining instantaneous reproductive number on a daily basis to effectively capture dynamic changes due to new detected cases and reduction of cases due to social distancing and nonpharmaceutical interventions. The prior value for R was selected at 2 and prior standard deviation of 5 to allow fluctuations in reporting of cases in the exponential phase. As there is limited access to epidemiological data of case, parametric with uncertainty (offset gamma) distributional estimate of serial interval was used. A mean of 2 and 7 days, with standard deviation of 1 was used to capture short and standard serial interval assumptions using 50 samplings of serial interval distribution. The Wallinga and Teunis method, as implemented by Ferguson^7^ is a likelihood-based estimation procedure that captures the temporal pattern of effective reproduction numbers from an observed epidemic curve. R was calculated using the web application EpiEstim App (https://shiny.dide.imperial.ac.uk/epiestim/)^7^. The descriptive statistics were used to compute mean and confidence intervals of the instantaneous reproductive number.

GENI score was anchored on the principle of rapid pathogen evolution between transmission events. This requires defining a suitable reference sequence of the outbreak, which is on the early stages the sequence nearest to the timepoint of the index case. For the case of COVID-19, the reference sequence is Wuhan seafood market pneumonia virus isolate Wuhan-Hu-1 NC_045512.2^16^. Publicly available virus sequences were retrieved from GISAID (supplementary Table 1) with whole genome variant determination using Snippy v4.6.0^17-19^. The average mutation per isolate was divided to the total epidemic curve days to derive a daily epidemic mutation rate and scaled to a monthly rate. We calculated the average nucleotide change per month to be 1.7 (95% CI 1.4-2.0), which was within boundaries of another estimate with the substitution rate of 0.9 × 10^−3^ (95% CI 0.5-1.4 × 10^−3^) substitutions per site per year^20^. We derived a transformed value of this rate before integrating it with epidemiological information. The output from the variant calling step was then used to determine GENI score by calculating the nucleotide difference. The basis for GENI score cutoffs to estimate transmission dates are derived from accepted evolutionary inference of mutation rates of COVID-19.

**Table 1.** Country-specific Instantaneous Reproductive Number (R) estimates for COVID-19 as of March 1, 2020.

|  |  | Instantaneous Reproductive Number (R)<br>with different serial intervals |  |
| --- | --- | --- | --- |
| Country | Cases | 2 days | 7 days |
| Mainland China | 79251 | 1.6 | 2.1 |
| South Korea | 3150 | 2.8 | 25.6 |
| Italy | 1128 | 8 | 57.0 |
| Iran | 593 | 2.8 | 17.1 |
| Japan | 241 | 3.6 | 2.2 |
| Singapore | 102 | 3.3 | 1.6 |
| France | 100 | 2.9 | 16.9 |
| Hong Kong | 95 | 2.6 | 1.6 |
| Germany | 79 | 3.1 | 17.2 |
| United States | 70 | 4.3 | 1.7 |
| Kuwait | 45 | 2.6 | 15.3 |
| Spain | 45 | 3.7 | 10.8 |
| Thailand | 42 | 3.8 | 1.7 |

We defined four epidemic curve (epicurves) stages to provide a clear method to define increases in the outbreak. The ‘index stage’ is characterized by the first report (index case) or limited local transmission indicated by intermittent zero incidence creating undulating epicurve. Secondly, which is distinctly different from stage 1, is the ‘takeoff stage’ in which the troughs are almost at same level of the previous peak and no longer touches zero, suggesting sustained local transmission. The ‘exponential stage’ is characterized by the classical hockey stick like sharp uptrend where the outbreak is moving quickly and large number of new cases are emerging. The last stage is ‘decline’ and is noted when the outbreak has reached the peak and cases being reported are lower than the peak, which will ultimately result in few to no new cases being reported, yet viral circulation is likely still occurring.

## Results

We determined the outbreak dynamics of pandemic COVID-19 by classifying each country’s status according to epicurve stages with a framework of a) index b) takeoff c) exponential d) decline as a clear method that can be used to benchmark metrics that include R and viral genome diversity. First, we calculated R using the instantaneous method using two serial intervals (2 and 7 days; Table 1). As of March 1, 2020, this framework defined global epicurves of COVID-19 outbreaks as gaining momentum globally with 52 countries were in the index stage. Three countries were in the exponential stage and five countries in the takeoff stage (Figure 1). China was the only country that reached the peak of the epicurve and characterized to be in the decline stage - decreasing cases. At this point there was no evidence of any other country near the decline stage and some countries that were poised to move into the takeoff and exponential phase.

**Figure 1.**
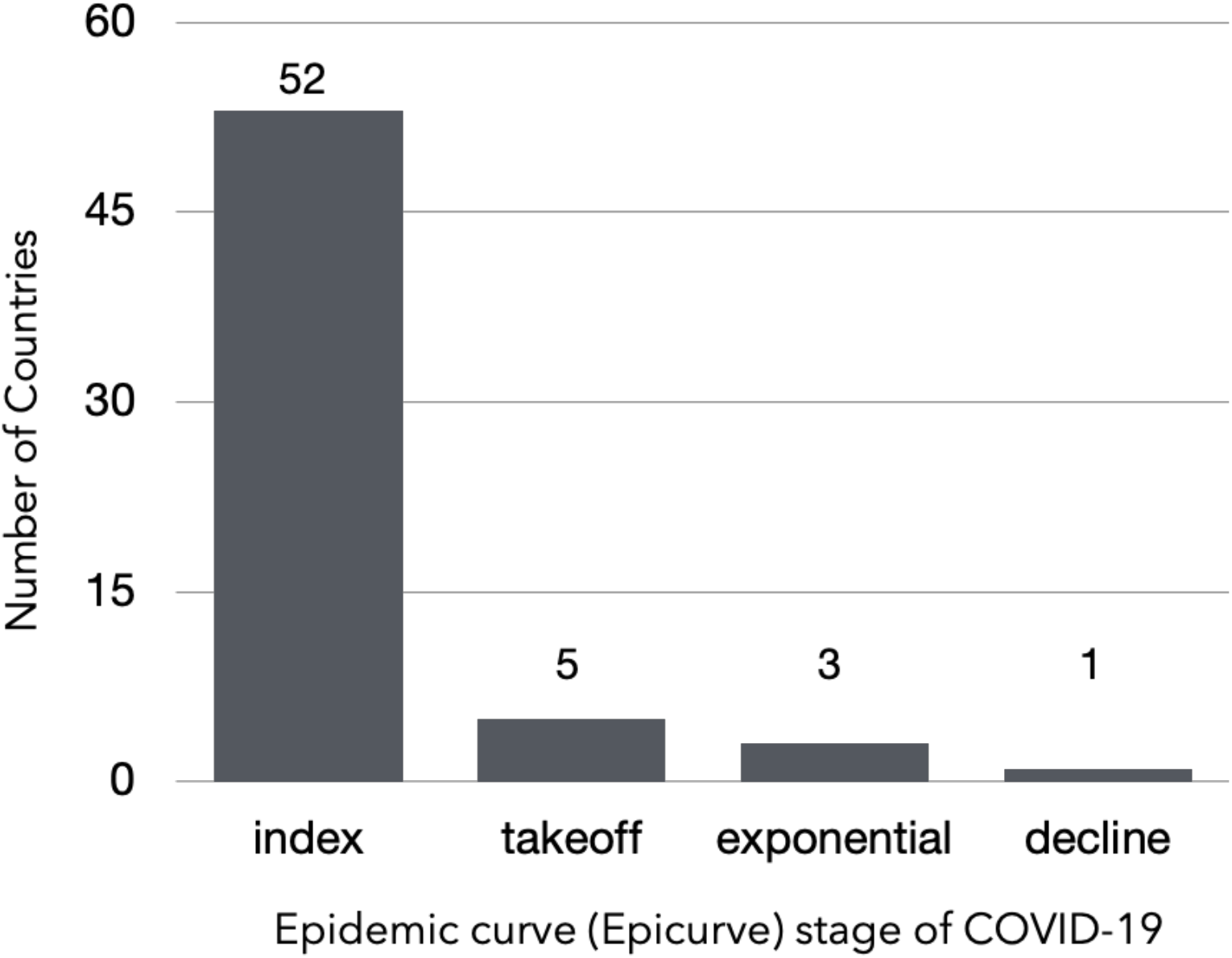
Distribution of country classification based on COVID-19 epicurve status.

Instantaneous R sensitively described real-time shifts of COVID-19 incidence captured within each epicurve stage (Figure 2). The decline stage in China was reflected by a decrease in R estimates in the latter stages the outbreak and relative to the early estimates: 1.6 (95 % CI 0.4-2.9) and 1.8 (95 % CI 1.0-2.7) for 2- and 7-days serial interval, respectively. Superspreading events inflated R estimates seen in exponential stage that was observed in South Korea: 2.8 (95% CI 0.6-5.3) and 25.6 (95 % CI 3.0-48.2) for 2- and 7-days serial interval, respectively. Efficient disease control was instituted in Singapore enabling it to remain in the index stage while Japan was moving to the takeoff stage characterized by increased R estimates 3.6 (95% CI 0.4-7.3) 2.2 (95% CI 1.3-3.0) for 2- and 7-days serial interval, respectively. The R estimates overlaps for all exemplar country outbreak stages in the two serial interval scenarios, suggesting that the transmission could be as short as 2 days. These estimates were relatively lower than previously reported, bringing to light possibility of transmission in the incubation period that is associated with rapidly expanding outbreaks, which is currently being observed in many European countries.

**Figure 2.**
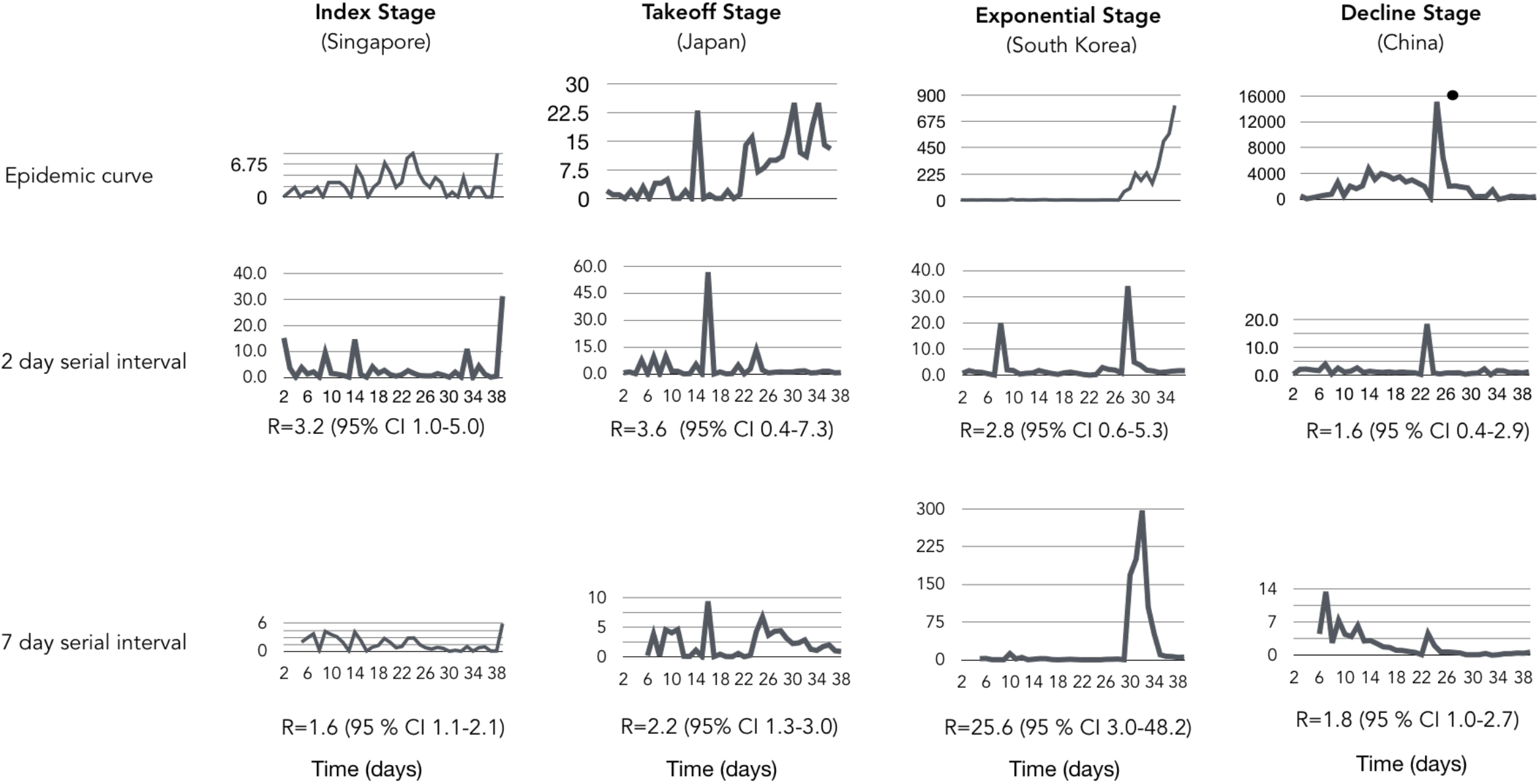
Instantaneous reproductive number estimates for different stages of the COVID-19 epidemic curve: a) index (Singapore) b) takeoff (Japan) c) Exponential (South Korea) d) decline (China) in short (2 days) and standard (7 days) serial interval. Decelerating stage of epidemic curve results to a reproductive number lower than 2 for both serial intervals, epidemic curve with multiple introductions yields 2-day serial interval with higher reproductive number and exponential serial interval yields higher reproductive number for the 7-day serial interval. Dot (.) the surge in the epidemic curve of China corresponds to the alteration of the case definition of COVID-19 by broadening confirmed cases with pneumonia confirmed with CT (computed tomography) scan. South Korea’s higher reproductive number is due to cryptic transmission associated with a secretive cult with altered health seeking behavior.

Low detection of COVID-19 was observed in representative countries in the index stage with low R values (<2) that can be attributed to effectiveness of social distancing intervention (i.e. Hong Kong) or under detection for countries with limited testing (i.e. United States) (Figure 3a). Sustained local transmission was occurring in five countries that were progressing into takeoff stage (Japan, Germany, Spain, Kuwait and France) as measured by R values (>2) (Figure 3b). The magnitude of spread was apparent with relatively higher R estimates (>10) in Italy, Iran and South Korea, which demonstrated sudden surges in incidence due to prior undetected clusters in part but other factors may contribute to this observation (Figure 3b). This significantly increased the instantaneous R estimates versus other methods of estimation but allows a more obvious depiction of the surge of cases allows differentiation of the takeoff stage from exponential stage.

**Figure 3.**
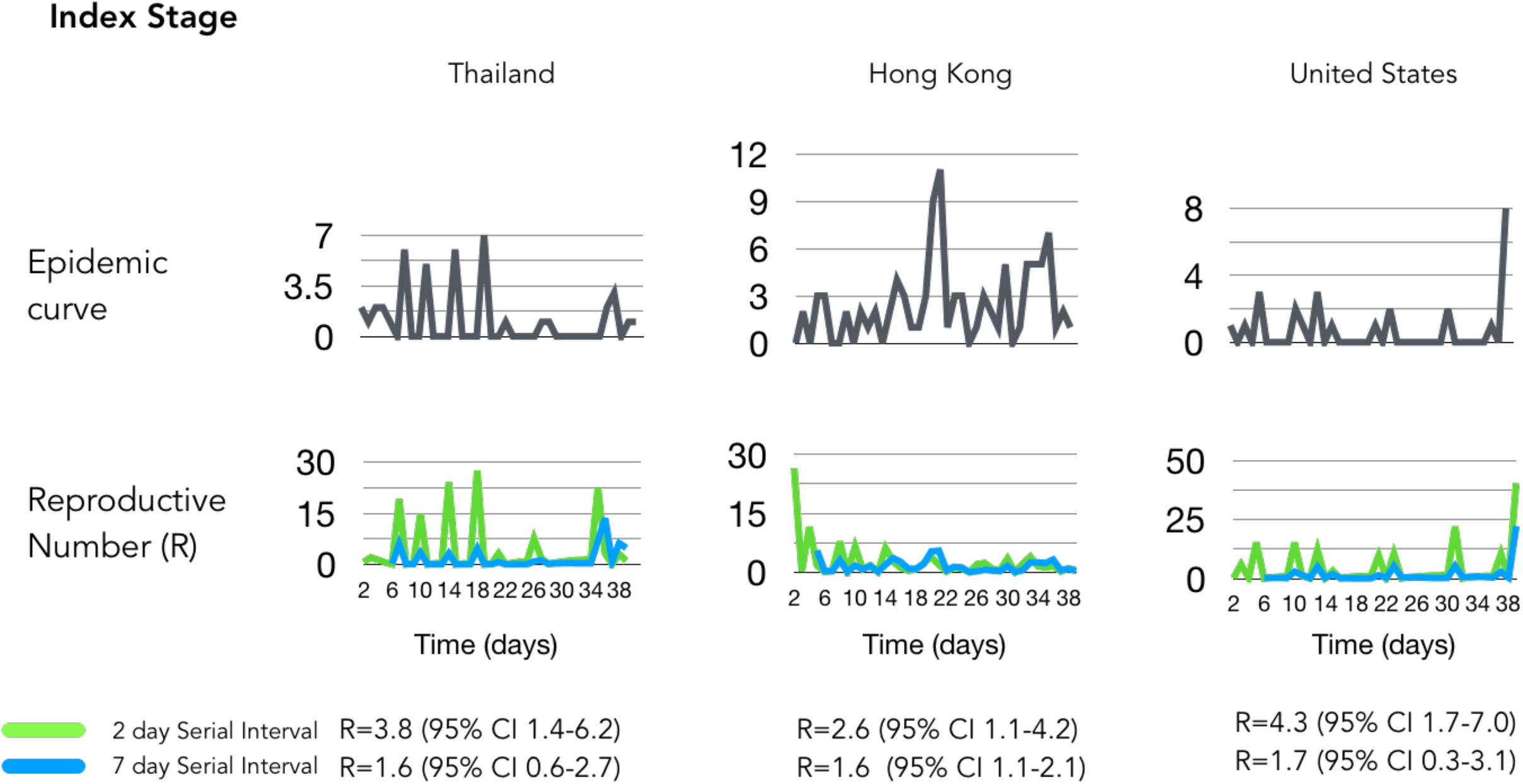
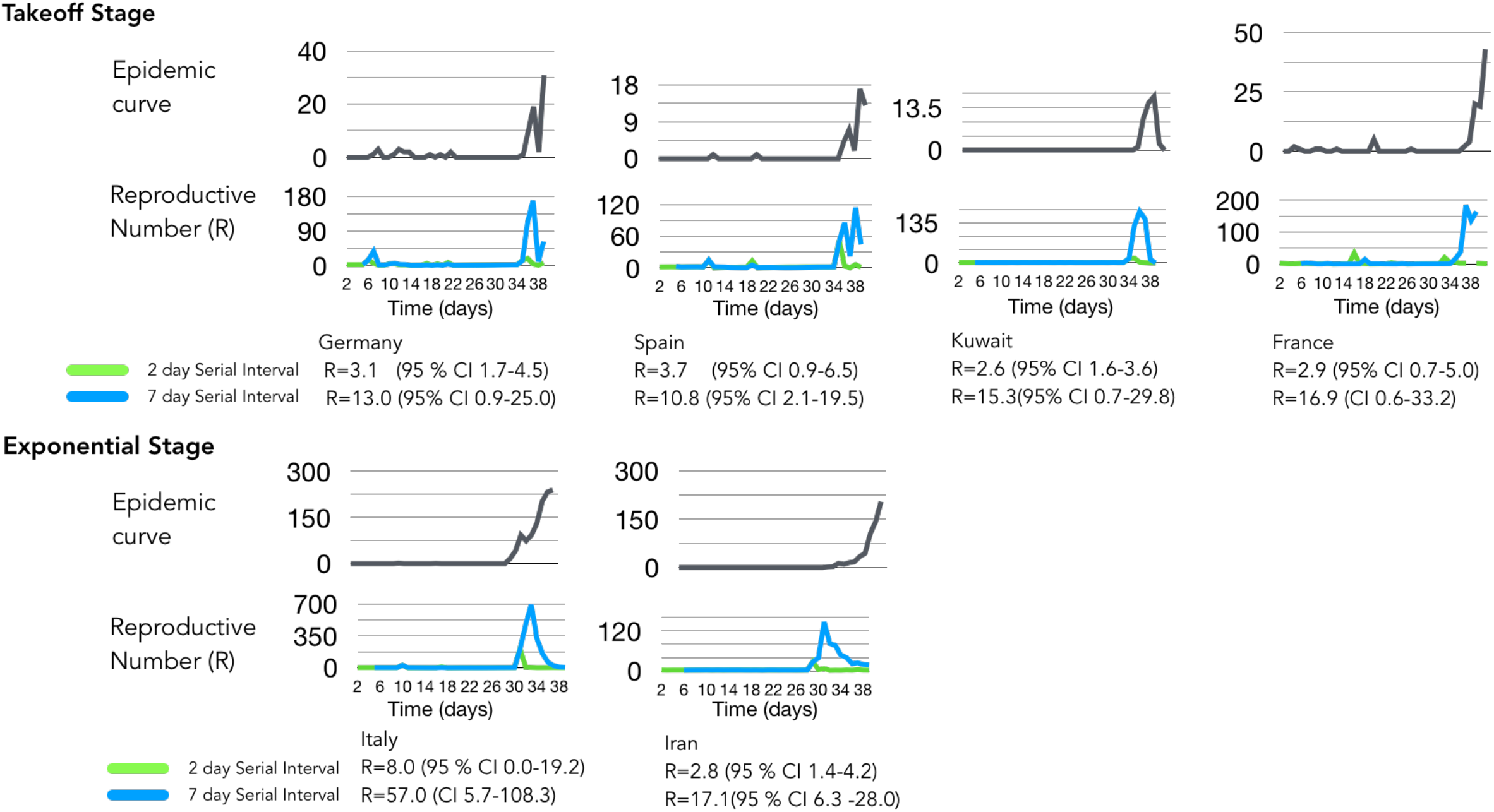
Epicurve estimates with different serial intervals. Panel A represents Epicurves and instantaneous R values for index stage countries using 2- and 7-day serial interval. Panel B Global dynamics of COVID-19 using instantaneous estimate of reproductive number with 2-day serial interval. Under preincubation period infectivity scenario, reproductive numbers globally increasing (> 2). Italy’s R = 8 is highest due to late detection of infection clusters. This higher R estimate is due to a huge bump in cases combined with diagnostic gap of low-level incidence. The same surge dynamics is seen in South Korea. Global dynamics of COVID-19 using instantaneous estimate of reproductive number with 7-day serial interval. Italy’s R value inflates to 57 with the 7-day serial interval assumption and overlaps with the lower threshold of 2 day serial interval R estimate. This estimation depicts a decreasing pattern for countries multiple introductions like Singapore, Hong Kong.

We further examined the value of computing country-specific instantaneous R by comparing different temperature range (tropical versus temperate) and population density. Population density of key cities (Table 2) and the higher temperature range values were used for selected countries; however, no direct link was observed. Increases in the South Korean outbreak was associated with a secretive religious group Shinsheonji (73% cases of COVID-19 in South Korea) located mainly in Daegu with a lower population density 883/km^2^ as compared to the rest of the areas with an outbreak^21^ and likely explain the outbreak expansion in the early epicurve. Religious beliefs that modify health seeking behavior particularly reporting clinical signs of COVID-19 combined with continued large group gathering prevented early detection of the outbreak. While most countries (Table 2) have cooler temperatures (10-6°C), Singapore’s temperature higher indicated that local transmission occurred at higher temperatures and suggests that temperature shifts will not likely change transmission. These commonly accepted environmental and behavioral activities did not explain the epicurve. This led to the hypothesis that the viral genomic variation underpinned changes in cases during outbreaks in each country.

**Table 2.** Epidemiological Parameters and instantaneous R estimates. The population density for South Korea is based on Daegu where 75% of the cases are reported.

|  | Reproductive Number (R) | Temperature (°C) during outbreak | Population Density (people/km<sup>2</sup>) | Interpretation in consideration of the epidemiological curve |
| --- | --- | --- | --- | --- |
| Singapore | 3.3 | 32 | 8136 | Imported cases, limited local transmission |
| France | 2.9 | 10 | 4300 | Imported, Local transmission >1-2 month |
| Italy | 8 | 10 | 7200 | Imported cases, Local transmission >1 month |
| United States | 4.3 | 9 | 8444 | Imported cases, Local transmission >2 month |
| South Korea | 2.8 | 6 | 883 | Imported cases, Local transmission >1-2 month |

**Table 3.** Relationship of Pathogen Genome Identity (GENI) Score derived from mutational difference from the index genome (Wuhan isolate of COVID-19 or cluster isolate reference from multiple outbreak regions outside of territory).

| Equivalent Pathogen Genome Identity (GENI) score for COVID-19 | Clinical Interpretation and Epidemiological Inference | Notes |
| --- | --- | --- |
| 0-2 | No difference from index case isolate genome or reference, imported case if there is no prior report, indicative of acute transmission <1 month | Reference genome is primarily earliest isolate available. |
| 3-4 | recent local transmission (average 1-2 months) if there are no prior report of cases | Subsequent outbreak clusters can serve as sources of introduction hence near neighbor reference has to be selected to generate an accurate GENI score. |
| >4 | sustained local transmission (greater than 2 months) if there is are no prior report of cases | Subsequent outbreak clusters can serve as sources of introduction hence near neighbor reference has to be selected to generate an accurate GENI score. |

We determined the relationship of epicurve stage with viral genetic variation using a metric that merges absolute genome variation with the rate of genome change to create the GENI metric that anchored population genome diversity with the rate of evolution for the SARS-Cov-2. To examine how the viral genome diversity was associated with the epicurve stages we first examined the index stage (Singapore) and the exponential (South Korea). Integration of GENI scores successfully distinguished the index from exponential stage (Figure 4). An increase in GENI scores was associated with exponential stage with a median score of 4, suggesting that the viral diversity and rate of mutation played was directly proportional to case increases during this stage. Singapore (index stage) effectively controlled the disease before becoming exponential had a GENI median score of 2. This was found in multiple time points during the outbreak were multiple mutation events were directly associated with increases in cases. While China is in the decline stage the retrospective association with R, cases, and GENI provided longitudinal evidence of multiple expansion in cases with mutation events in the viral genome, especially early in the epicurve. The repeated viral mutations and epicurve expansion were associated in each time point over 3 months, in three countries, and in three different outbreak stages. This finding is useful in integrating virus genome diversity and evolution into assessment of outbreak status in an outbreak between countries but also within the epicurve when combined into a triad with instantaneous R estimates. The proportionality of GENI scores with the epicurve stage indicates its value in determining the outbreak status and the importance of generating population scale genome sequence resources.

**Figure 4.**
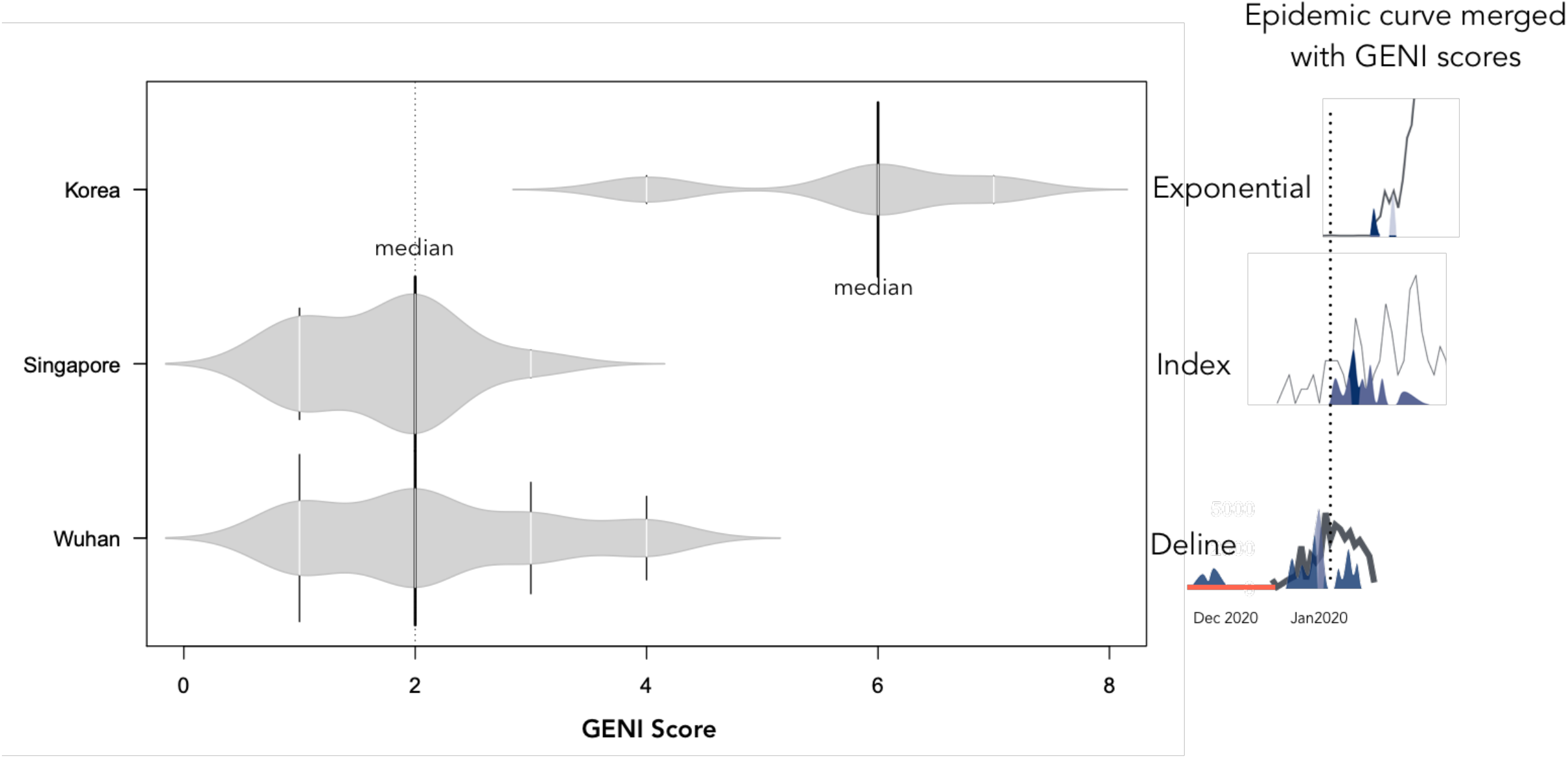
Relationship of pathogen genome identity (GENI) score with the temporal signal along the epidemic curve. Local transmission is captured by virus mutation as expressed in GENI score values. GENI scores of SARS-COV2 isolates are relative to Wuhan reference strain Wuhan-Hu-1 NC_045512.2. The red line in the China epicurve represents the time before an outbreak was determined yet genome sequences were circulating. The blue shaded curves indicate GENE scores directly overlaid with the outbreak curve. The dotted line represents the common point in time as a reference for visualization. The GENI score and epicurve show similarity except in China as the outbreak advanced to takeoff and exponential the GENI score increased while in the index stage example of Singapore the outbreak was contained and the GENI score remained <2.

A framework to merge epidemiology and population genomics was derived from this study as a systematic method for molecular epidemiology (Fig. 5). It requires dynamic measurements be taken for R and longitudinal efforts to determine each virus whole genome sequence. Using this triad of measurements accurately and quickly provided insight to measure outbreak progress but also provides an evidence-based method for interventions. This study demonstrated an advancement of how to use population genomics in a viral situation where the mutation rate is fast and the genome diversity of the population is extraordinarily high. GENI provided a missing method that defines how to use viral genome mutation dynamics and genome population diversity, which is only observable using large numbers of genomes, that occurs during an outbreak.

**Figure 5.**
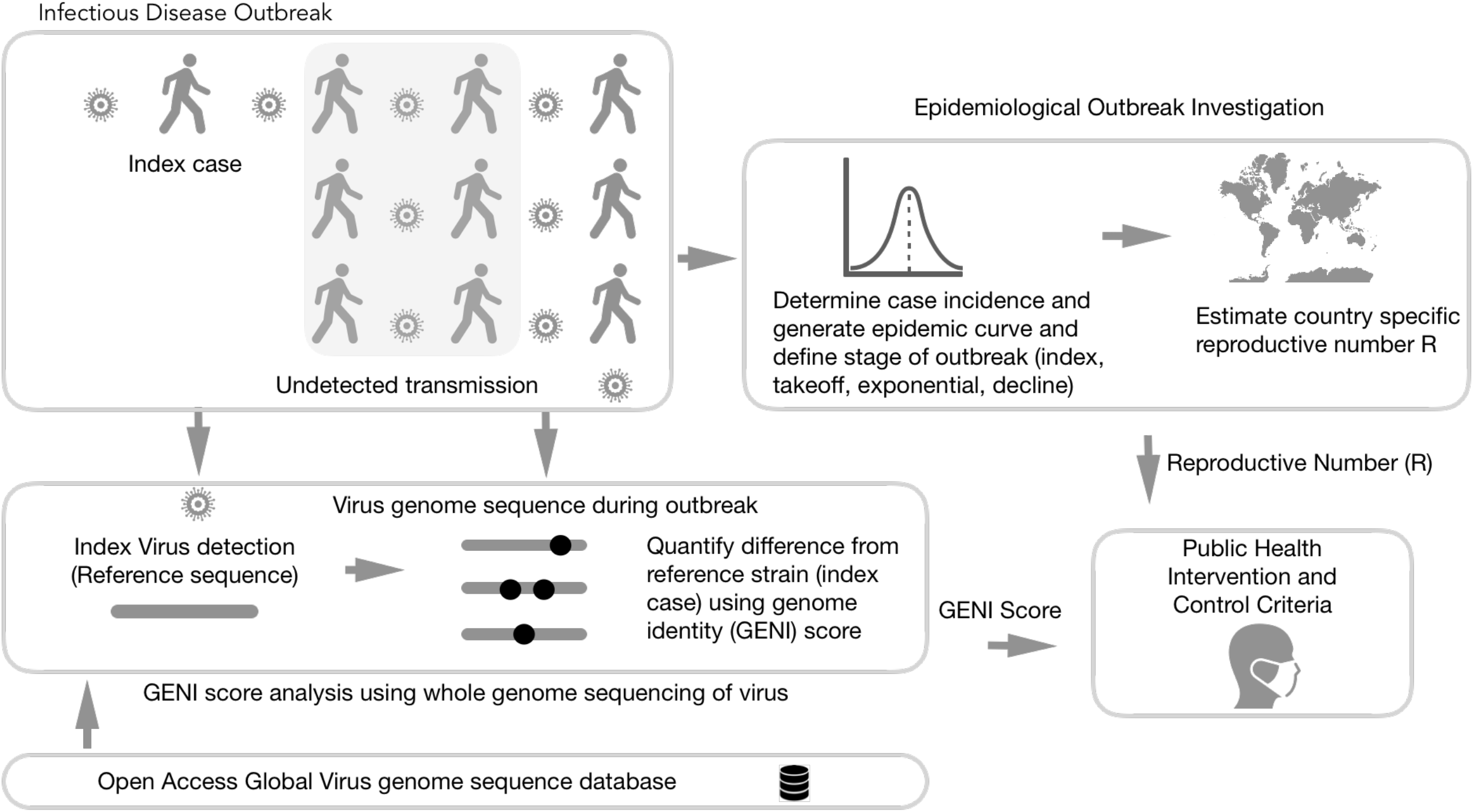
Integration of genomic and classical epidemiology for outbreak investigation. The foundation of epidemiology is the accurate and timely reporting of cases which enable the calculation of the number. Genomic Identity (GENI) score is formulated from genomic data of pathogens to differentiate imported cases versus local transmission and measure time of cryptic spread. Together these two epidemic values deliver insight that can be directly used for making decision criteria for public health intervention.

## Discussion

Public health response is proportional to the severity and transmission dynamics of an infectious disease outbreak. This requires epidemiological metrics that can be used as decision criteria, and ideally, they can be used to assess impact of the intervention. In this work we determined that R is much more dynamic in the COVID-19 pandemic than previously appreciated by country as well as over the outbreak within each country (Fig 2-3). The instantaneous R estimation with a serial interval of 2 was extremely sensitive to shifts in the epicurve during the index phase (Fig 2-3). Singapore is an excellent example of effectively controlling and containing the COVID-19 outbreak. They previously designated a response system called Dorscon (Disease Outbreak Response System Condition)^22^ providing a systematic approach to control so that they have not moved past the index phase. In contrast, most other countries in this phase are poised to move into the takeoff phase (Fig 3). The transition into the takeoff phase signified a transition from a 2-day serial interval to a 7-day serial interval that was more sensitive to shifts in the epicurve.

While estimates of R alone is insightful in retrospect, gaps in epidemiological surveillance due to several factors creates blind spots that hindered the ability to determine interventions. To overcome this limitation, we merged GENI estimates based on whole genome sequence variation and mutation rate with the epicurve and R and provided a predictive triad of measurement that resulted in insight that accurately refined case expansion (Fig. 4). Each phase of the outbreak was characterized with mutations that led to new cases in established outbreaks by case definition. The merged information indicate that China found variation in the viral sequence much earlier than the outbreak cases increased. Independent of the phase framework merging sequence variants with the epicurve found that new cases were observed in the same timeframe as new sequence variants were found. Previous studies that the relationship of genomic diversity with epidemic severity (i.e. R) found no clear link^20^. However, by merging instantaneous R, the epicurve stage, and the GENI index it is clear that a link exists for each country examined that resulted in a direct link between outbreak dynamics and the absolute genomic mutation with the mutation rate. The GENI index provides a basis to examine imported cases or locally spreading, both of which addressed this current work using established metric - R and novel integration of viral whole genome sequences to define changes in the sequence that are directly linked to increases in cases. This leads to an epidemiological metric that is scientifically robust and at the same time can convey complex biological properties to enable an efficient characterization of an outbreak in combination. Transforming complex pathogen characteristics was made usable to public health and medical field using the GENI score as a complete merged information set with other characteristics of the outbreak.

Previous outbreaks, such as Ebola, employed state of the art analysis using phylodynamics that is anchored on the genetic evolution^13^. Inference such as time to most recent common ancestor allowed estimation of outbreak origin, population size, and R – yet this was not integrated into the outbreak dynamics and stage of advancement in the outbreak. This type of analysis is possible because genomic sequences carry temporal signals and when used in context with sample from different timepoints, previous divergence can be determined. The GENI score includes these signals and expands their use by merging them with the outbreak dynamic using the population genome variation as well as the mutation rate.

This inherit information is not limited to viruses. Another recent example in a bacterial setting was the cholerae outbreak in Haiti wherein the phylogenetic analysis resolved the origin of the pathogen^23^. However, for this analysis to succeed, a substantial database of genome sequences is needed, collected across time and geographic location to enable placement in a phylogenetic context. As outbreaks as bound to happen in the future, investment in cataloguing the genomic space of pathogens is as ever important^24,25^. It is critical to obtain COVID-19 sequences from humans as well as other animals that have zoonotic potential, as was demonstrated previously with zoonotic *Campylobacter* species^26,27^. Creating sequence repositories of pathogens is critical and underway for various pathogens^25^ as well as COVID-19^18^.

Prior work forewarned the practice of being overly dependent on early estimates of R alone^28^. By having the most accurate possible information for a dynamic metric and taking into account the complex dynamics that factor in the calculation of R along with merging this the genomics of the pathogen is a robust and insightful method to assess outbreak dynamics, as demonstrated in this study. Openness and data sharing of incidence reports and sequences at unprecedented scale is being done in this pandemic and it is paying rewards^29^. Leveraging on these resources opens unexpected collaboration and avenues for applying relevant bioinformatic and disease modelling skills across the scientific community to solve global public health problems. Examples that hindered this were observed in several countries that led to cryptic spread of the disease in countries. Additionally, lacking the epidemiological infrastructure and genome sequencing capabilities limit this approach that is not acceptable for modern public health. However, without the appropriate technical skills in the performing complicated phylogenetic inference, utility of such innovation will be limited. Establishing a protocol for merging epidemiology and genomics was defined in this work (Fig. 5) and can be instituted globally.

## Conclusion

This study integrated population genomics into epidemiological methods to provide a framework for molecular epidemiology. Specifically, this study demonstrated using epicurves, instantaneous R estimates, and GENI specific case increases in COVID-19 are directly associated with viral mutation. It was demonstrated that the pandemic is poised to become larger and that mutation will be associated with the increase in cases. Exemplar outbreaks, such as Singapore, found increases in cases with viral mutations that were effectively controlled. However, other outbreaks had expanding R estimates during the outbreak, as well as numerous viral mutation events. Use of epicurve stages, instantaneous R estimates, and GENI provided a robust and accurate framework to monitor outbreak progression to different stages with direct association between cases and increases in each metric.

## Data Availability

the data are public and a list of the sequences used is included

## Acknowledgement

We gratefully acknowledge the authors, originating and submitting laboratories of the sequences from GISAID’s COVID-19 Genome Database. We also thank the global community for rapid information sharing that enabled integration of these data.

## Notes

### Competing Interest Statement

The authors have declared no competing interest.

